# Improved user-friendliness in the design and analysis of FAVITES-Lite simulations

**DOI:** 10.1101/2024.06.10.24308702

**Authors:** Kate Stadler, Kayla Phillips, Niema Moshiri

## Abstract

After the rise of several different simulation tools that can simulate different aspects of an epidemic, FAVITES provides a framework to simulate an end-to-end epidemic (from contact network generation to infection transmission to viral phylogenies). To increase accessibility, FAVITES-Lite was developed with the key functionalities of FAVITES with the sacrifice of some flexibility. It remains that configuring and analyzing a complicated simulation like FAVITES or FAVITES-Lite can be extremely daunting, so we created example configuration files to help users learn the process behind parameter choice and a graph-visualization web application to help users visualize the results of their simulations.

## Introduction

Several tools have been developed to model different aspects of an epidemic, including contact networks, infection transmission, viral evolution and more. Epidemiological parameters such as transmission rate can be estimated using a maximum-likelihood model based on phylogenetic trees, and transmission patterns can be described throughout the tree (Stadler & Bonhoeffer, 2013). Using pathogen DNA and collection dates, *outbreaker* can reconstruct transmission trees and gain insight into super spreaders, undetected cases, and separate introductions (Jombart et al., 2014). Prevention methods were also evaluated by simulating an HIV-1 epidemic based on demographics, sexual partnerships, viral introductions, HIV infection, transmission, acquisition, Antiretroviral Therapy (ART) status, intervention, sequence sampling, ancestral HIV relationships, and sequence evolution (Ratmann et al., 2016).

FAVITES was introduced in 2019, building off of previous models to provide a framework to model epidemics fully end-to-end (social contact network, transmission history, incomplete sampling, viral phylogeny, error-free sequences and real world sequencing imperfections) (Moshiri et al., 2019). Throughout each module in FAVITES, users can choose models and parameters for those models while existing tools at the time simulated only a subset of each step. FAVITES-Lite was introduced to simplify the process of setting up and running the simulation. FAVITES-Lite incorporates the key functionalities of FAVITES, which comes at the cost of some flexibility. It also contains scripts to visualize the simulated data that is output at its completion, producing a graph image of the current number of individuals in each state over the course of the simulation. In an attempt to make this specific script more user-friendly, we created a more interactive experience through a web application that allows the user to see exactly how many individuals are in each state at any time of the simulation.

For each of the modules, users must choose a model and model parameters. However, these model and parameter selections must be structured as a “configuration file” in the JSON format, which can be difficult for users to structure manually. In another attempt to increase ease of useability, a configuration designer web application (https://niema.net/FAVITES-Lite) was introduced to help users choose these models and parameters and automatically convert them to a JSON file format, to be used when running the simulation. The web application offers model descriptions and limited parameter descriptions (Fig. 1).

**Figure 1:**
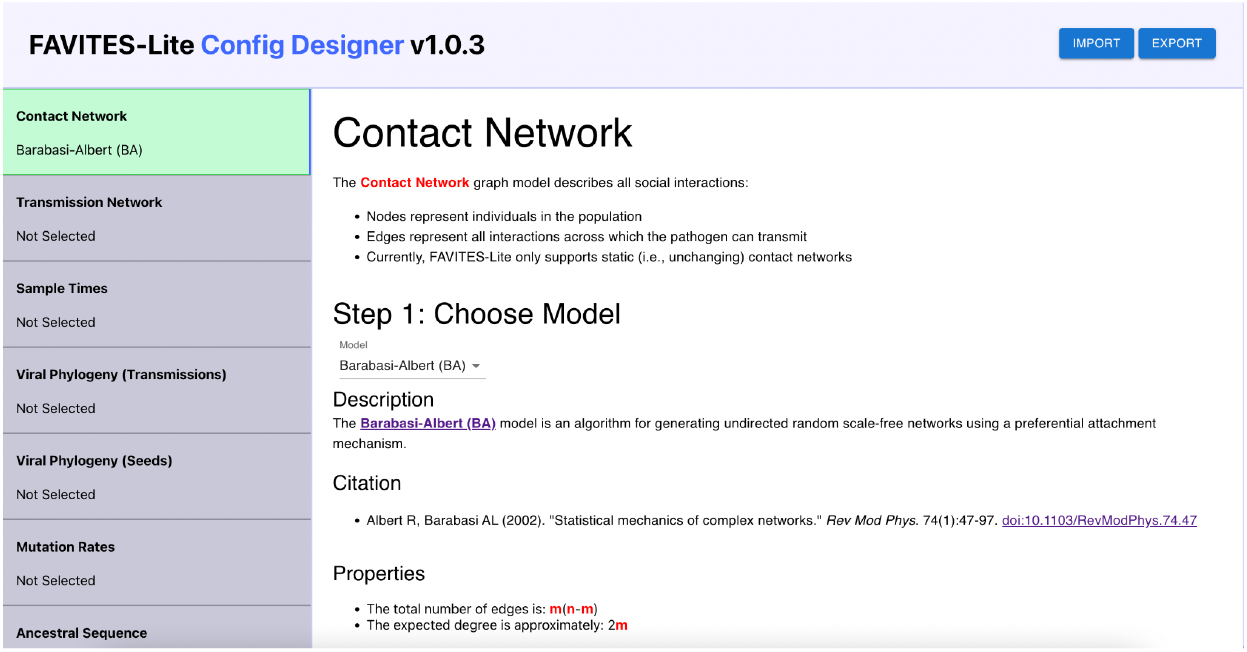
Screenshot of the FAVITES-Lite Config Designer web application.

**Figure 2:**
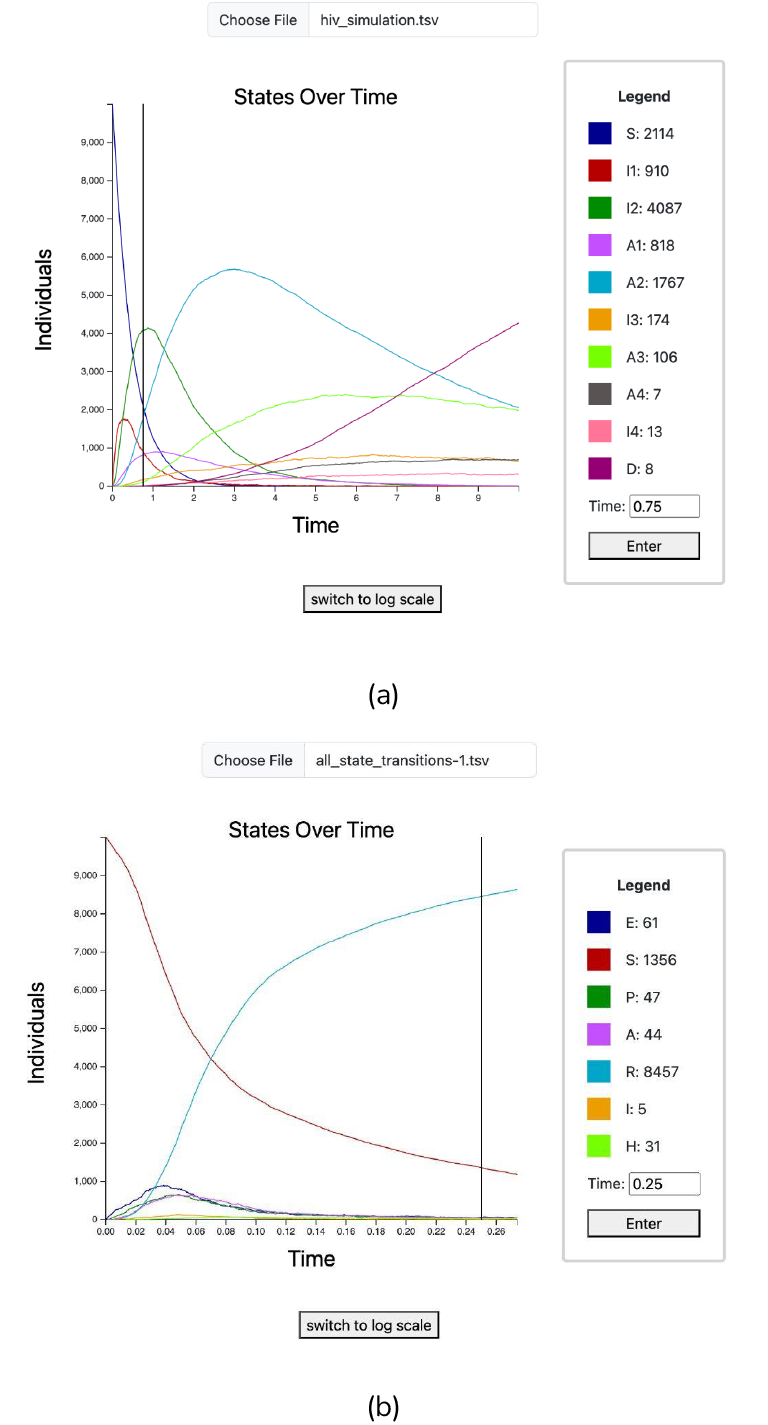
Screenshot of the States Over Time web application visualizing data produced by (a) a simulation modeling the HIV epidemic in San Diego County using the models and parameters described in Table 1, and (b) a simulation modeling the early SARS-CoV-2 epidemic in Wuhan using the models and parameters described in Table 2.

While this web application makes *inputting* each simulation choice significantly easier and more efficient, simply *choosing* models can be a complex task, and determining realistic values for each model parameter can be challenging. In this work, we aim to simplify the design and analysis of epidemic simulations by providing simple, context-rooted example configuration files, with each parameter choice explained (De Angelis et al., 2015). These example configuration files will provide the user with a starting point from which to design their own epidemic simulation experiment.

## Methods

### Epidemic-specific Example Configuration Files

Our goal was to create 2 example FAVITES-Lite configuration files: one modeling an HIV epidemic, and one modeling a SARS-CoV-2 epidemic. We started with three papers with an interest in their use of FAVITES. The first paper, Moshiri et al. (2019), used FAVITES to simulate HIV epidemics in San Diego and Uganda, comparing simulated data to real data. Sensitivity to parameters, parameter choices (such as time to stop ART), and clustering methods were also tested. The next two papers Pekar et al. (2021) and Pekar et al. (2022) used FAVITES to look at the timing and zoonotic origins of the SARS-CoV-2 epidemic, respectively. Beginning with a target time, place, disease, and population, we examined these papers for their parameter choice and reasoning. We also reviewed the papers for our chosen Transition Network models, including Granich et al. (2008) and Hao et al. (2020) when further specificity was needed. Finally, we used Zingoni et al. (2019) for information on HIV disease progression among Antiretroviral Therapy (ART) patients.

#### HIV-1 epidemic among MSM in San Diego County, 2016 through 2025

This configuration file is meant to define a simulation of transmission among MSM in San Diego, and includes modules for Contact Network, Transmission Network, Sampling, Viral Phylogeny, Mutation Rates, Ancestral Sequence, and Sequence Evolution. Only susceptible individuals were chosen to be modeled, so non-susceptible individuals were left out of this simulation. Many of our models and parameters were taken from Moshiri et al. (2019), which used FAVITES to compare simulated and real HIV datasets in San Diego and Uganda.

1. For the contact network, we chose the Barabasi-Albert (BA) network model, emulating the simulation from Moshiri et al. (2019). Because BA networks have power-law degree distributions, it is suitable for social and sexual contact networks, so we chose to root our configuration file in the context of the MSM community.
2. For the transmission network, we used the transmission model proposed by Granich et al. (2009), which has 11 states (Non-Susceptible, Susceptible, 4 HIV stages that can each be Treated vs. Untreated, and Deceased), and 46 parameters (Table 1). We chose to include 10,000 individuals in our network in order to limit computing time. One individual was initially infected with HIV stage one, and the rest started off susceptible. For the transition rate from susceptible to infected stage 1, we calculated the expected value of new HIV diagnoses out of a population of 10,000 susceptible individuals in the San Diego MSM community, to be used as a fixed transition rate in our fixed Transmission Network. We used an MSM population estimate of 80,968 in San Diego 2016 (Grey et al. 2016) and a count of 321 diagnoses in San Diego in 2016 with reported MSM male-male sexual contact, or MMSC (County of San Diego HIV/AIDS Epidemiology Report—2016). By dividing the number of diagnoses by the estimated susceptible population (in our case, the MSM population in San Diego), and multiplying by our desired number of individuals for our network, we estimated (321/80,968) * 10,000 = 39.645 diagnoses in San Diego in 2016 as a result of MMSC, in a pool of 10,000 susceptible individuals. Transition rates are the reciprocal of the expected time to next arrival, which is equal to the expected value. Infectivity was pulled from the supplementary material table S4 in the original FAVITES paper. Transition rates between states and mortality and mortality rates on ART were motivated by Zingoni et al. (2019), which studied HIV disease progression among Antiretroviral Therapy (ART) patients in Zimbabwe estimated mean sojourn time and total length of stay in each stage of HIV while adhering to ART. Note that, while we expect social dynamics of HIV transmission to differ between Zimbabwe and San Diego County, the parameters that were motivated by Zingoni et al. (2019) only pertain to progression of individuals throughout the different stages of HIV, which is a biological/physiological phenomenon, *not* parameters related to diagnosis rates or adherence to ART, which would be largely impacted by social, behavioral, and economic factors.
3. Each individual was sampled when they first started ART (Moshiri et al., 2021).
4. A Non-Homogenous Yule tree with rate function exp(-t^2^) + 1 was chosen to describe the topology and branch lengths of the viral phylogeny of the seed individuals, which was pulled from the supplementary material in Moshiri et al. (2019). The Non-Homogeneous Yule tree was chosen because with the rate function, it could model the short internal branches close to the root of a real HIV tree. The tree was then scaled so that the height matched the 1980 time-of-most-recent ancestor.
5. Mutation rates were sampled from a truncated normal distribution because other distributions deviated significantly from real ones. Location and scale parameters for the truncated normal distribution were pulled from the parameters used in the San Diego simulations run in the original FAVITES paper.
6. Ancestral base frequencies were based on exact base frequencies, which was chosen as a simple option with the ability to specify the frequency of each base. The length of the ancestral sequence is 9200, matching the length of the HIV genome. Frequencies of each base were calculated from the NCBI HIV-1 reference sequence (NC_001802.1).
7. Sequences were evolved using the General Time-Reversible (GTR) + Gamma model, which matches the model used in the original FAVITES paper. The base frequencies, base transition rates, and Gamma shape parameter were pulled from the Supplementary Material in the original FAVITES paper.

#### SARS-CoV-2 epidemic in Wuhan, late 2019 through early 2020

Most of our models and parameters were pulled from Pekar et al. (2021), which used FAVITES to investigate the timing of the SARS-CoV-2 index case in Hubei province.

**Table 1:**
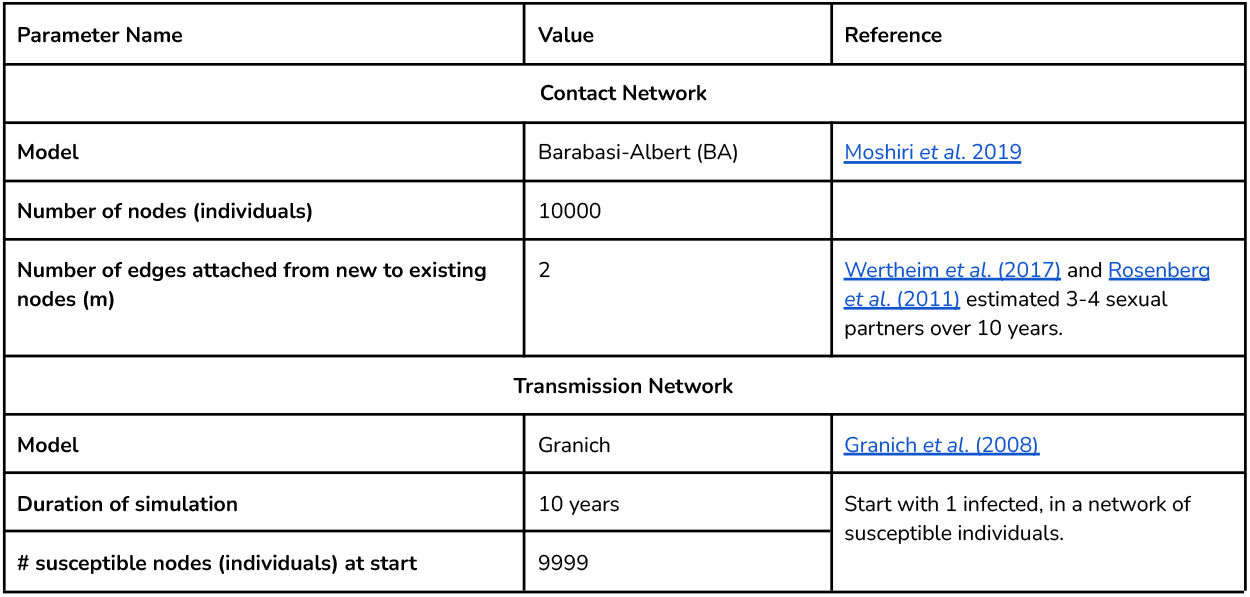
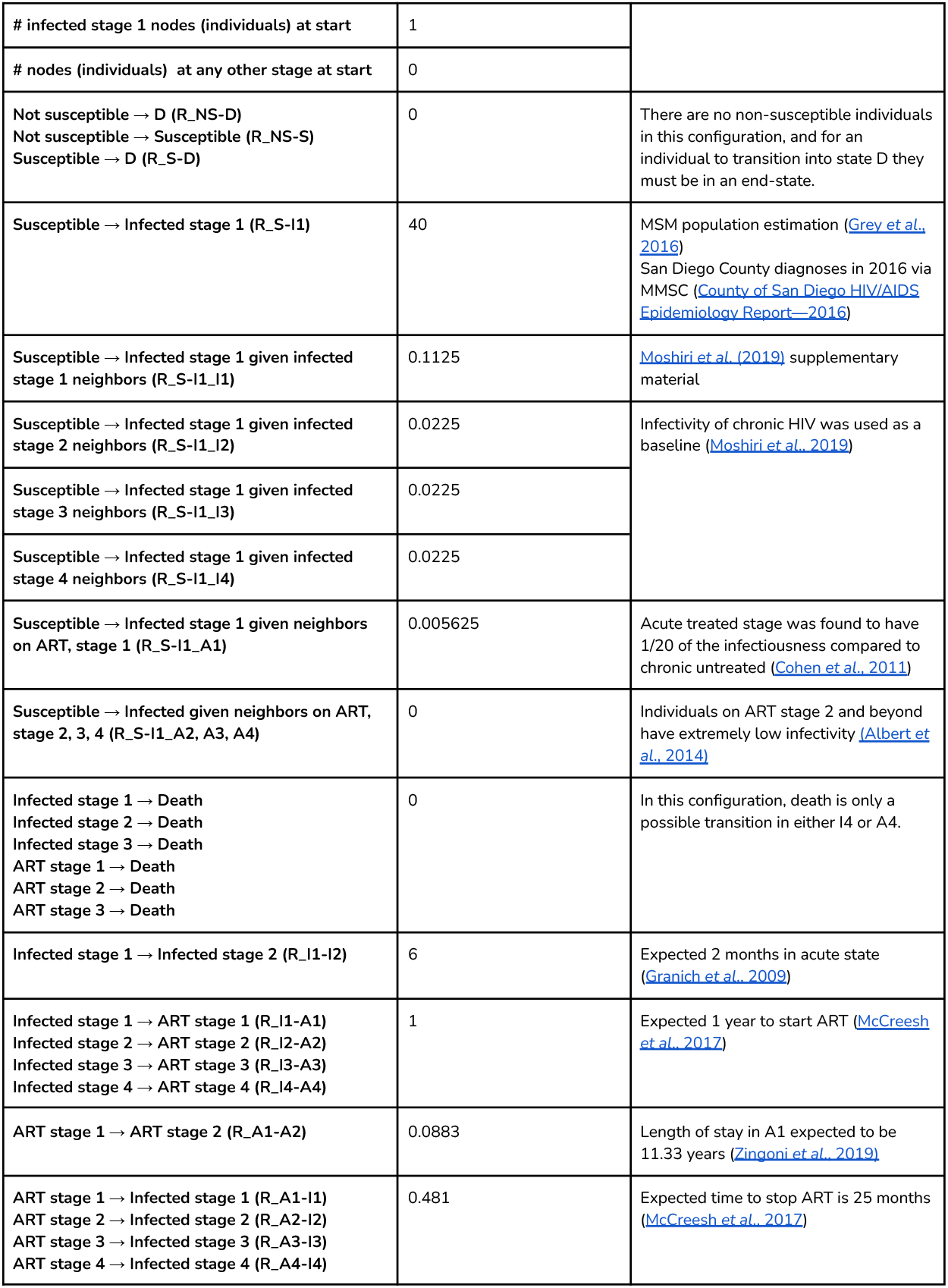
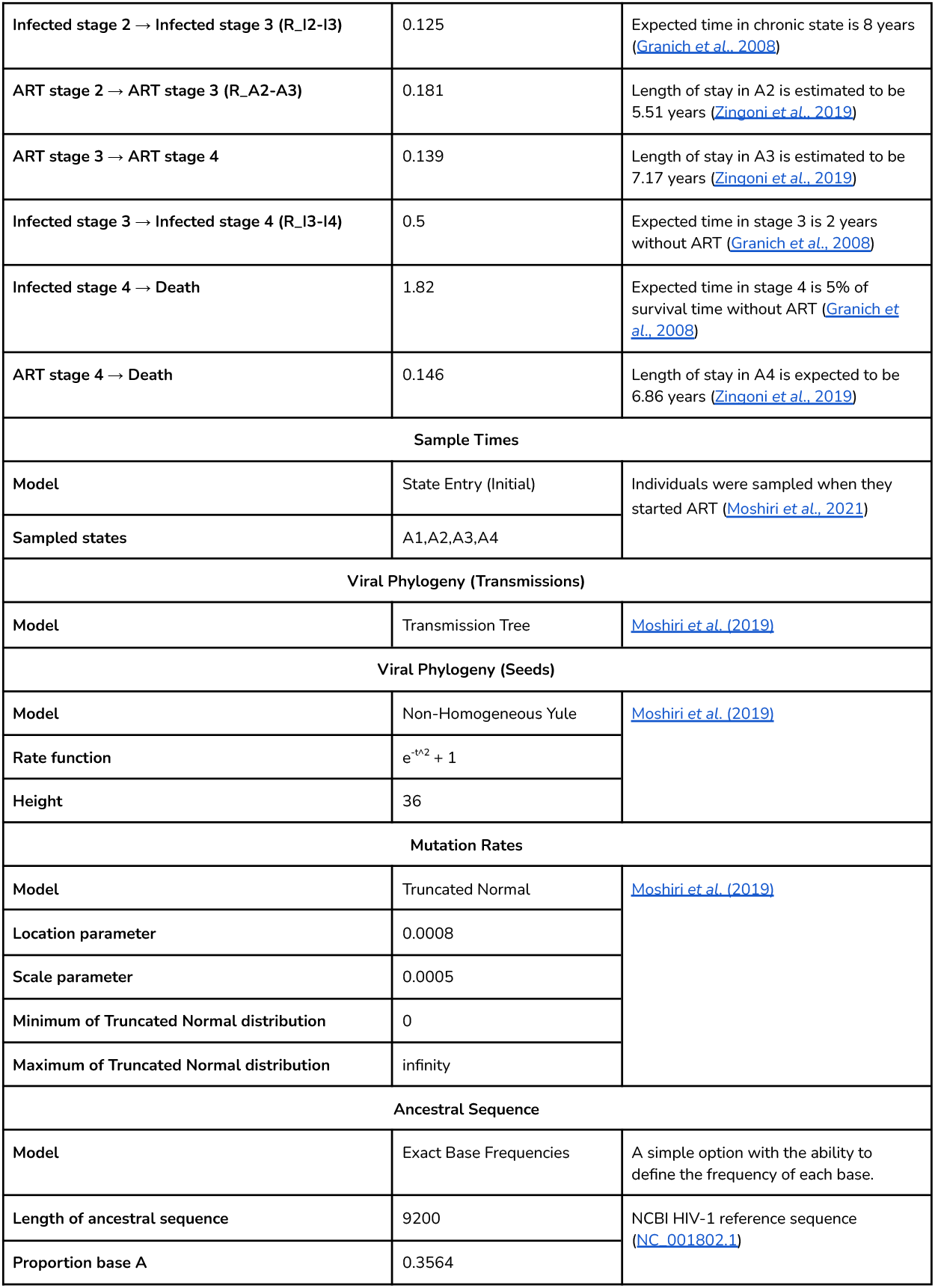
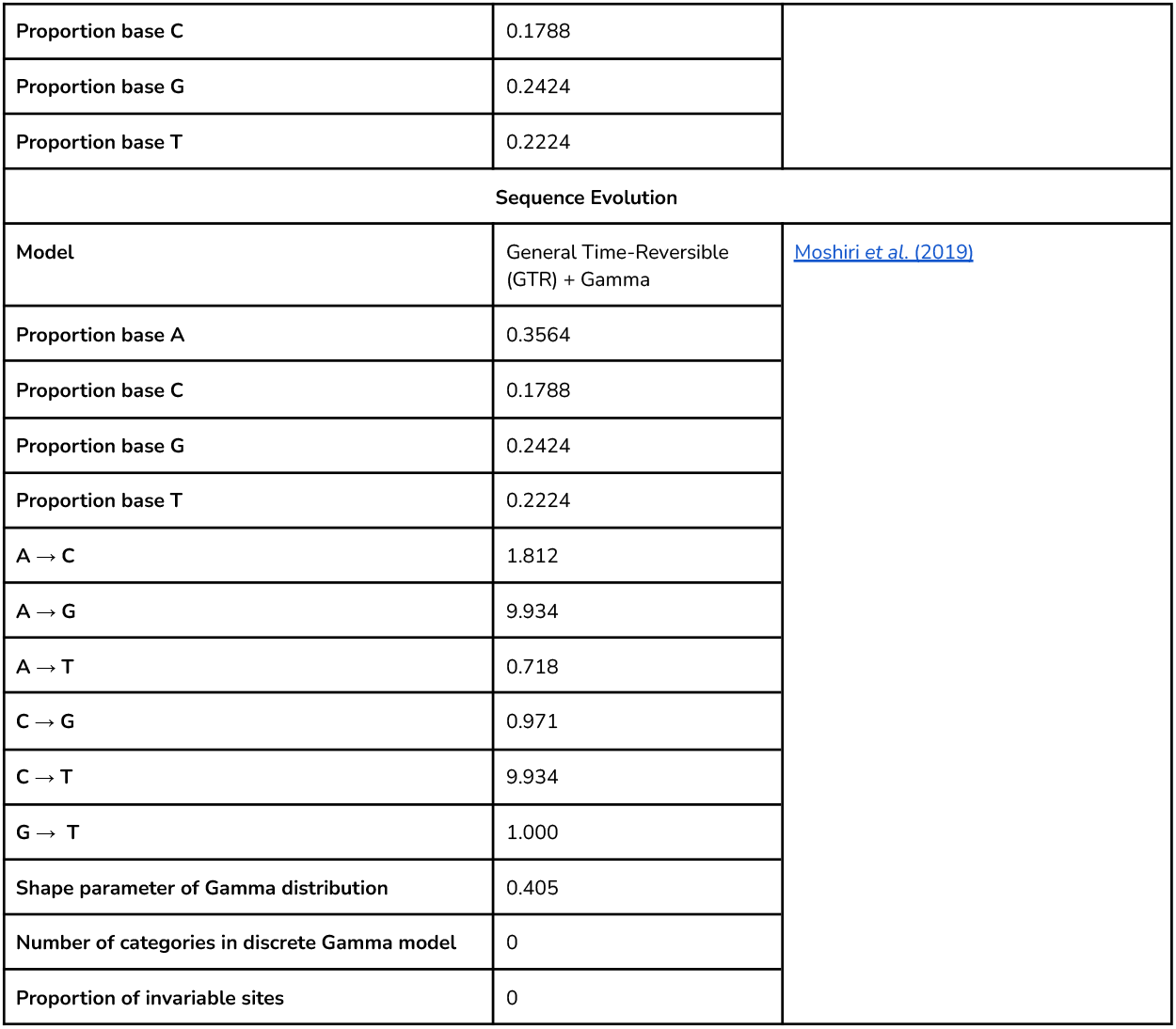
HIV-1 epidemic among MSM in San Diego County, 2016–2025.

**Table 2:**
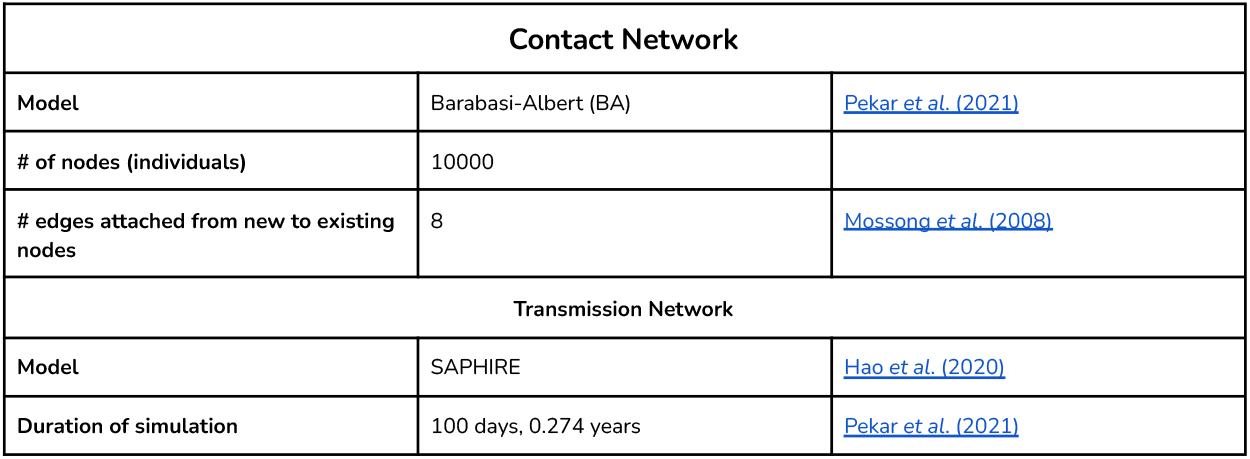
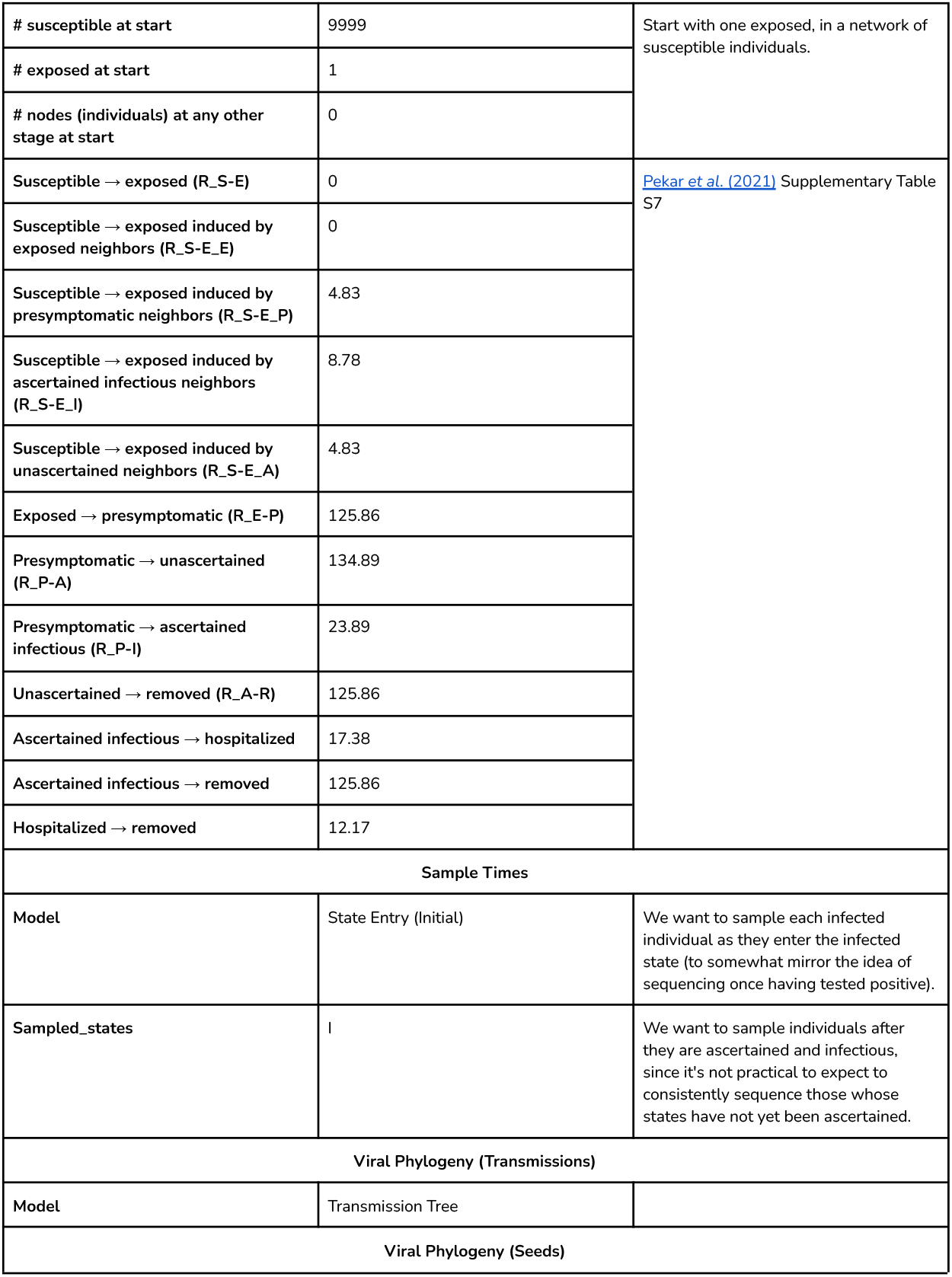
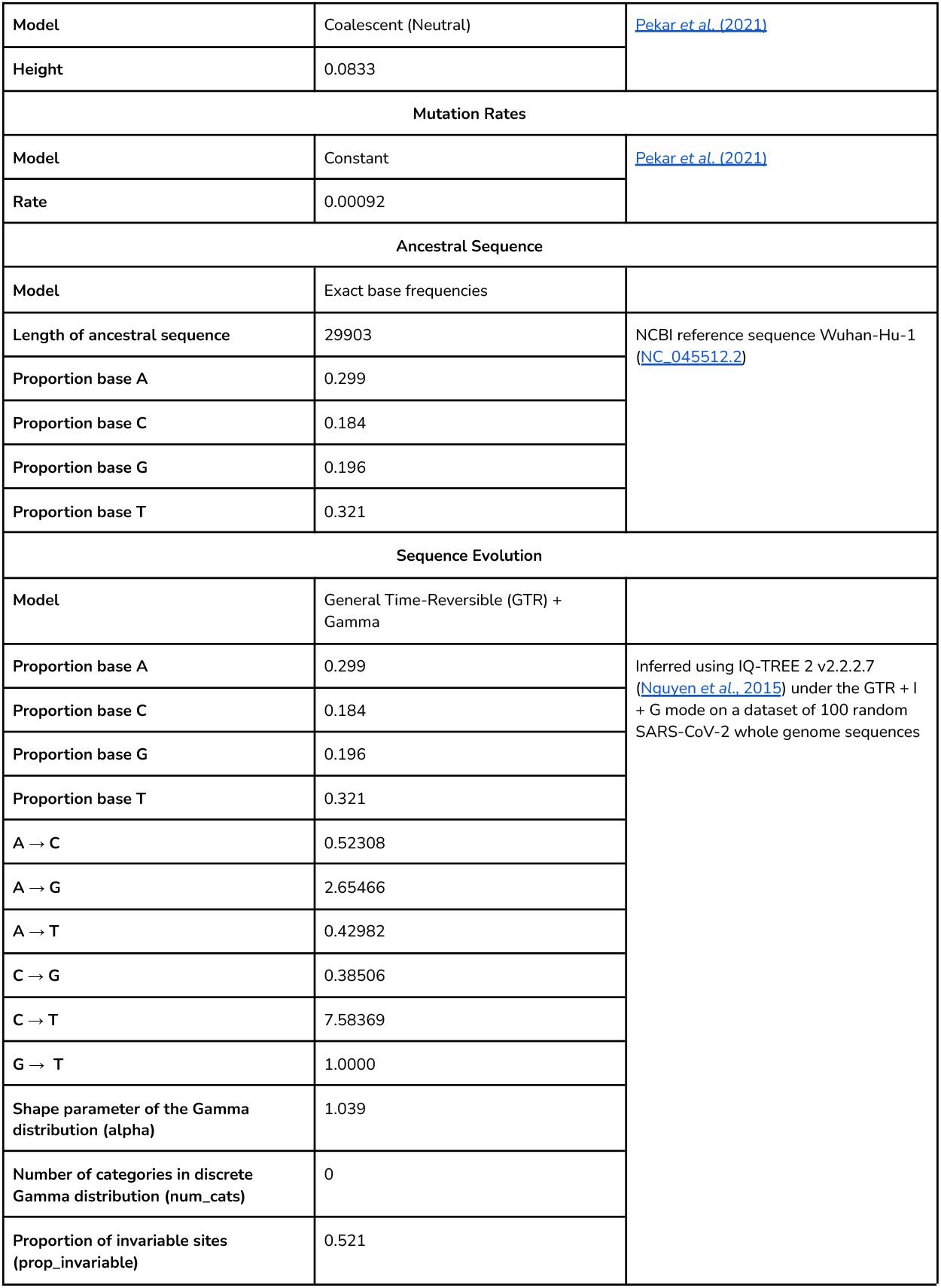
SARS-CoV-2 epidemic in Wuhan, late 2019 – early 2020.

1. The BA model was chosen because its scale free properties “recapitulate infectious disease spread” (Pekar et al. 2021). A quantity of 10,000 individuals were chosen for ease in running the simulation. 8 edges were chosen to be attached from new to existing individuals based on an average value of 16 contacts per day (Mossong et al. 2008). Note that the BA model produces networks with relatively wide tails, and networks simulated with 10,000 individuals and an expected degree 16 will have a handful of individuals with upwards of ∼500 contacts (i.e., ∼5% of the total population).
2. The SAPHIRE model was chosen to illustrate the dynamics of COVID-19 transmission (Hao et al. 2019). Duration was chosen to be 100 days because the simulation is meant to look at the start of the COVID-19 pandemic. 1 individual started as exposed and the rest were susceptible. Transition rates between states were pulled from the supplementary material in table S7 in the COVID paper.
3. The state entry (initial) model was chosen for sampling so that we could sample each individual once they became ascertained-infected, reflecting the idea that individuals would get sequenced once they officially tested positive.
4. A Transmission Tree model was chosen to describe the topology and branch lengths of the viral phylogeny so that coalescent events occur as late in time as possible.
5. The Coalescent (neutral) model was chosen to model variation in DNA sequences due to genetic drift and/or mutation.
6. Based on the results of an analysis using BEAST (Drummond & Rambaut, 2007) from Pekar et al. (2021), mutation rates were constant at a rate of 0.00092, inferred from BEAST.
7. Exact base frequencies were used for Ancestral Sequences. The length of the genome was chosen to be 29903, the length of the COVID-19 genome, and frequencies were calculated from the NCBI reference sequence Wuhan-Hu-1 (NC_045512.2).
8. Like the HIV configuration file, the GTR + Gamma model was chosen. Parameters were inferred by running IQ-TREE 2 v2.2.2.7 (Nguyen *et al*., 2015) under the GTR + I + G mode on a dataset of 100 random SARS-CoV-2 whole genome sequences produced by the Andersen Lab (https://github.com/andersen-lab/HCoV-19-Genomics).

### States-over-Time Web Application

The web application requires the input of the TSV file, one of the output files from a FAVITES-Lite simulation named “all_state_transitions.tsv” by default. The web application was built using HTML, CSS, and JavaScript. The javascript library d3.js was used to visualize the data. The web application is deployed using GitHub Pages.

## Results

### Epidemic-specific Example Configuration Files

#### States-over-Time Web Application

The web application aims to help intuitively and interactively visualize the number of individuals in each transmission state over time, processing raw epidemic simulation data that are otherwise difficult to comprehend. The web application accepts a tab-delimited (TSV) file containing all state transitions generated by running a FAVITES-Lite simulation. The web application requires no prerequisite setup or installation: users can simply navigate to the web application using any modern web browser on any modern operating system.

The web application then generates a lineplot visualizing the number of individuals in each state (vertical axis) over time (horizontal axis). The bounds of the axes are automatically scaled to the input dataset’s bounds, and the vertical axis can be toggled between linear and logarithmic scale with the click of a button. Colors for the curves of each state are automatically generated to be maximally separated for ease of interpretation of the plot. A legend is automatically produced to the right of the graphs, corresponding the colors of the curves on the graph to the states they represent.

To enable closer inspection of the epidemic states over time, the graph also has an interactive vertical bar that displays the exact number of individuals in each state at any given time of the simulation. The number of individuals for each state at the specified time are shown in the legend, to the right of the colors and states. These values automatically update whenever the specified time is changed. To choose a time, the user can either type a specific time into the input box at the bottom of the legend, or the user can hover over the graph at the desired time. To fix the vertical bar’s position (i.e., the time for which the states’ counts are displayed), the user can simply click on the graph, which will prevent mouse interactions from changing the selected time. To resume mouse interaction, the user can click the graph again to unfix the vertical bar’s position and allow the selected time to move freely again.

## Discussion

The parameters and explanations in these files were meant to help a first-time user start with ballpark figures that can help them learn about the process of choosing these parameters. There are several limitations to these files, but these shortcomings can actually be helpful in designing future simulations or adjusting current ones.

First, the context of these files is very specific: the parameters chosen for a San Diego HIV simulation in 2018 would be far different from a San Diego HIV simulation in 1990 or a Miami HIV simulation in 2018. Difference in demographics would change the contact network and/or transmission network setup. Users should not necessarily use these files as dictionaries for their own simulations—they should use them as a practical way to get an example of how their parameters might be chosen. The references included can also be a resource for much more information.

But even with the freedom to choose 40+ parameters, flexibility and nuance are balanced against complication and useability, as is the nature of parameterization. FAVITES-Lite currently offers eight choices of static networks, each of which may suit a different context well. For example, we chose the Barabasi-Albert (BA) model for both of our configuration files because we wanted a power-law degree distribution, and beyond the number of nodes, the BA model requires just one free parameter which controls the degree distribution. However, this model produces networks with relatively wide tails, leading to a handful of individuals with upwards of ∼500 contacts in our 10,000 individual simulation (*m* = 8 ⟶ expected degree = 16). A more complex model that can better control the degree distribution may be more appropriate to accurately model contact networks, but it would consequently be more difficult to parameterize. While we aim to capture reality as much as possible, we only aim to provide new users a *starting* point with reasonable models and model parameters in order to strike a balance between realism and simplicity, and we leave further efforts to calibrate simulations to better fit specific real-world scenarios of interest to users as they design their simulation experiments.

PATH 4.0 (Singh et al., 2021) was developed as an agent-based evolving network, with infected persons and their immediate contacts as agents. When new people become infected, the challenge is to decide which individual (degree, risk group, age, location) should be added to the network. PATH 4.0 uses a changing probability of transmission per act, modeled as a function of disease, care stage, risk group, number of partners, and age. Changes in sexual behavior are also updated for each time step. Even the connections between individuals have attributes: partnerships have features initiation age, initiation time, termination age, and termination time. This model is able to capture many of the specific features that make up a contact network—with the contact networks offered in FAVITES-Lite, individuals have no attributes other than state, and edges don’t have features that would model some type of partnership. PATH 4.0 is also dynamic, allowing for changes in the size of the network and the risk of transmission over time. PATH 4.0 models total prevalence, diagnosed prevalence, annual incidence, and annual diagnoses, distributed by risk group and age. It can also generate clusters, meant to be similar to those detected by nucleotide sequence data, through its transmission network. While PATH 4.0 offers an extremely detailed and dynamic transmission network and contact network model, it does not simulate viral phylogeny or sequence evolution, which FAVITES-Lite can do. This is another example of the tradeoff between the ability to model an aspect of an epidemic, introducing complexity to increase the accuracy of the model, and the ability to model an end-to-end epidemic, introducing complexity to model many dependent phases of a simulation.

Though it is unable to capture many details than other, more niche simulations, FAVITES-Lite is unique in that it aims to simulate the full end-to-end epidemic. In and of itself, it is a way to increase usability by simplifying different aspects of a pandemic into palatable modules. Within its framework, new models could be added to increase flexibility, but as always this would widen the scope of possible parameters even more. While the configuration files created gave some good example configurations for COVID and HIV, there are many more parameters and models that were not covered. Future developments could include a guide to all the models in the framework and their parameters, or more configuration files for different contexts.

While the configuration files provide the foundation for setting up the simulation’s parameters and models, the web application offers a user-friendly interface for visualizing and interacting with the simulation results. The web application is designed to provide more precise information about the number of individuals in each state at all times of the simulation.

However, there are additional features that could be added that would make the user experience more interactive. The user is able to input a time from the simulation, and the hover bar will snap to that location on the graph as the legend displays the number of individuals in each state at that time. To improve the web application, providing the user with more statistics about the data, such as the rates of change for each state or a derivative graph of the data, would be beneficial. These statistics could be used to analyze and predict trends in disease/mortality rates (Fay et al., 2006).

## Data Availability

All data are available online at: https://github.com/niemasd/FAVITES-Lite

## Acknowledgements

We thank Bradley Voytek, Natasha Martin, Ravi Goyal, Michelle Truong, Hyeri You, and Tara Tenenbaum for fruitful conversations.

